# Comparing DXA and MRI body composition measurements in cross-sectional and longitudinal cohorts

**DOI:** 10.1101/2024.12.12.24318943

**Authors:** Nicolas Basty, Marjola Thanaj, Brandon Whitcher, Jimmy D Bell, E Louise Thomas

## Abstract

In this study we assessed the accuracy of dual X-ray absorptiometry (DXA) compared to magnetic resonance imaging (MRI) for evaluating body composition. Using data from 32,961 participants in the UK Biobank, including 1,928 re-scanned participants after about two and a half years, we examined cross-sectional and longitudinal agreements in DXA and MRI measurements within android and gynoid regions. Our results showed that DXA reliably captured fat measurements but overestimated lean mass compared to MRI, particularly in android regions for men (4.10 vs. 1.74 kg) and women (2.92 vs. 1.10 kg). Longitudinal MRI data revealed a 4-5% muscle and lean mass decrease, undetected by DXA, which showed lean mass increases in women at the follow-up visit. Although DXA is practical for population-level fat assessments, MRI remains the preferred method for detailed and precise longitudinal body composition analysis.

## Introduction

Detailed body composition assessment, including the measurement of adipose tissue and muscle content and distribution, provides a valuable role for understanding trends in health and disease prevalence in large-scale population studies. They allow the development of targeted interventions to promote healthy ageing and mitigate the impact of conditions associated with an adverse body composition^1,2^. Additionally, these measurements are crucial for assessing the effects of lifestyle factors, such as diet and exercise, on disease mitigation and overall well-being. Longitudinal body composition monitoring is particularly significant in ageing populations since fat distribution and muscle mass changes are associated with sarcopenia and reduced functional capacity^3^.

Many methodologies have been developed to measure body composition, from indirect and accessible techniques such as body mass index (BMI), waist-to-hip ratio or skinfold anthropometry, to more direct but more expensive and time-consuming imaging-based methods including dual X-ray absorptiometry (DXA), computed tomography (CT) and magnetic resonance imaging (MRI) which have the advantage of assessing the distribution of adipose tissue and lean mass (or muscle).

DXA, initially developed for the assessment of bone mineral density (BMD), using low-energy x-ray acquisitions, was subsequently upgraded for the automatic derivation of overall adipose tissue and lean mass, as well as from relevant anatomical areas, including the android and gynoid regions^4,5^. Despite the planar nature of the acquired images, DXA provides estimates for the volume or mass of the body in three dimensions, offering a more comprehensive understanding of body composition compared with indirect methods. DXA is characterised by rapid processing, low levels of ionising radiation and relatively low cost, making it widely used for individuals and large-scale population studies. However, DXA has limitations, unlike CT and MRI, it cannot directly measure specific individual adipose tissue depots. DXA also provides an estimate of “lean tissue”, but it is important to note that so-called lean tissue includes other non-fat tissues besides muscle. Therefore, beyond appendicular measures DXA may not offer specificity for muscle tissue. Despite these limitations, DXA remains a widely used tool in body composition assessments, offering a balance of accuracy and practicality for various clinical and research applications^6^.

In contrast, MRI is generally regarded as the gold standard for assessing body composition. Its non-invasive and non-ionising three-dimensional approach allows volumetric assessment of organs, adipose tissue, and muscles, from the largest tissues down to the smallest organ substructures with millimetre precision. While MRI provides unparalleled detail and accuracy, its widespread use has been limited by cost, accessibility, and the time and expertise required for image acquisition and quantitative analysis. A trade-off between DXA and MRI depends on the specific research or clinical goals, balancing factors such as precision, cost-effectiveness, and practicality in the given context.

Although DXA is widely accepted for assessing body composition^,7^, and most studies demonstrate strong and significant correlations between MRI and DXA measurements^8,9^, discrepancies between the two methods have frequently been noted. DXA is reported to overestimate subcutaneous adipose tissue (SAT)^10^ and underestimate visceral adipose tissue compared to MRI, especially in individuals with low levels of visceral adipose tissue (VAT)^10^. Interestingly, Neeland et al noted that in addition to underestimating VAT at low VAT levels, DXA also modestly overestimated VAT at higher VAT levels. Similarly, although good correlations have been reported between muscle mass measurements by DXA vs MRI and CT^10–12^, the agreement between measurements is poor, with DXA overestimating lean or muscle tissue^12,13,14^. A potential reason for the lack of agreement may relate to the few direct comparisons using the DXA manufacturer’s predefined regions of interest against MRI equivalents.

Similar issues have been observed comparing DXA against MRI for measuring body composition in longitudinal studies. Here a confounding factor is the sample size of longitudinal comparison studies, which tend to be much smaller than those used in some cross-sectional studies. Studies focussed on body fat content in adults and children have demonstrated that DXA substantially underestimated longitudinal changes in VAT^15^, with no association between longitudinal changes in DXA vs MRI-VAT^16^. For lean mass and muscle, some studies in adults have reported comparable age-related loss of muscle mass by DXA and MRI^13^, while others have suggested that compared with MRI DXA is only sensitive enough to measure large changes in lean mass^17^. Similarly, studies in children are conflicting; Bridge et al reported age-related changes in both fat and muscle mass to be similar in children for both MRI and DXA^18^, while Lee and Kuk reported that DXA overestimated fat loss and muscle gain compared with MRI in a weight loss intervention (N=39)^19^.

To overcome these discrepancies approaches including deriving equations to convert DXA values to MRI equivalents^20^, or constructing linear regression models^21^ have been proposed. Whether these strategies would enhance agreement between methods in longitudinal studies remains uncertain. Thus, a more comprehensive study comparing DXA and MRI, using anatomically equivalent anatomical regions and a larger cohort of subjects, is clearly needed.

This study aimed to leverage the extensive UK Biobank data to evaluate the agreement between DXA and MRI measurements of adipose tissue, lean mass, and muscle mass, using comparable DXA-defined android and gynoid regions in the MRI analyses. Additionally, the longitudinal data from this large cohort allowed us to assess the effectiveness of DXA, relative to MRI, in tracking changes in body composition over time.

## Methods

### Image Acquisitions

MRI scans were acquired using a Siemens Aera 1.5T scanner (Syngo MR D13) (Siemens, Erlangen, Germany), with full details of the UK Biobank abdominal MRI protocol reported previously^22^. Our analysis used the neck-to-knee chemical-shift encoded MRI, commonly known as Dixon MRI. All data were processed and segmented using automated methods^23^, with the outputs arising from this work already returned and available from the UK Biobank (Category 158).

DXA scans were acquired using a GE iDXA (GE-Lunar, Madison, WI). Full details of the UK Biobank DXA protocol have been previously reported^22^. Fields available from the UK Biobank DXA scanner outputs include: category 124, “*Body composition by DXA*”^1^, specifically: *Android fat mass* (field 23245), *Android lean mass* (23246), *Android tissue fat percentage* (23247), *Visceral adipose tissue volume* (23289), *Gynoid fat mass* (23262), *Gynoid lean mass* (23263), and *Gynoid tissue fat percentage* (23264). The only DXA reported volume (in litres) is for VAT, all other measurements are provided as a percentage or mass (kg).

### Study Design and Cohort

DXA and MRI scans were acquired from every participant on the same day with no more than 2-4 hours between scans. Longitudinal data was acquired from a subcohort (n=3,088) after an average time interval of 2.3 ± 0.1 years, utilising the same scanners and protocols for each imaging modality.

Dixon MRI data for 41,707 participants that was acquired at the baseline imaging visit between 2014 and 2020 with data comprising imaging, health-related diagnoses and biological measurements were available. Participants with missing body composition measurements by DXA were excluded from the study. More specifically, android region measurements for 10,815 participants were missing VAT volume measured by DXA from the baseline imaging visit and for 1,011 participants from the follow-up imaging visit. For the gynoid region, 10,674 participants were missing lean and fat mass measured by DXA from the baseline imaging visit and 978 participants from the follow-up imaging visit. Out of 3,091 UK Biobank participants who underwent a follow-up imaging visit^24^, 2,906 participants had complete imaging data at both visits and were included in the study. Overall, after these exclusions, a dataset of 32,787 (30,892 from baseline imaging visit and 1,895 from follow-up imaging visit) and 32,961 (31,033 from baseline imaging visit and 1,928 from follow-up imaging visit) participants with android and gynoid measurements, respectively, were included in the final analysis.

### Data Analysis

To generate a like-for-like comparison between DXA and MRI measurements, we reproduced, on the MRI scans, the DXA regions as defined in the software user manual^2^, which states that “*the android region of interest (ROI) has its lower boundary at the ‘pelvis cut’ and its upper boundary to lie at 20% of the distance between the pelvis and the ‘neck cut’. The gynoid ROI is to be twice the height of the android ROI, and the upper boundary to be below the pelvis cut line, 1*.*5 times the height of the android ROI”*. Therefore, we defined the android and gynoid regions in the MRI utilising the same anatomical landmarks (Figure 1), with the segmented pelvic bones used as *the* “*pelvic cut*” and the top of the MRI acquisition as the “*neck cut*”^23^.

**Figure 1.**
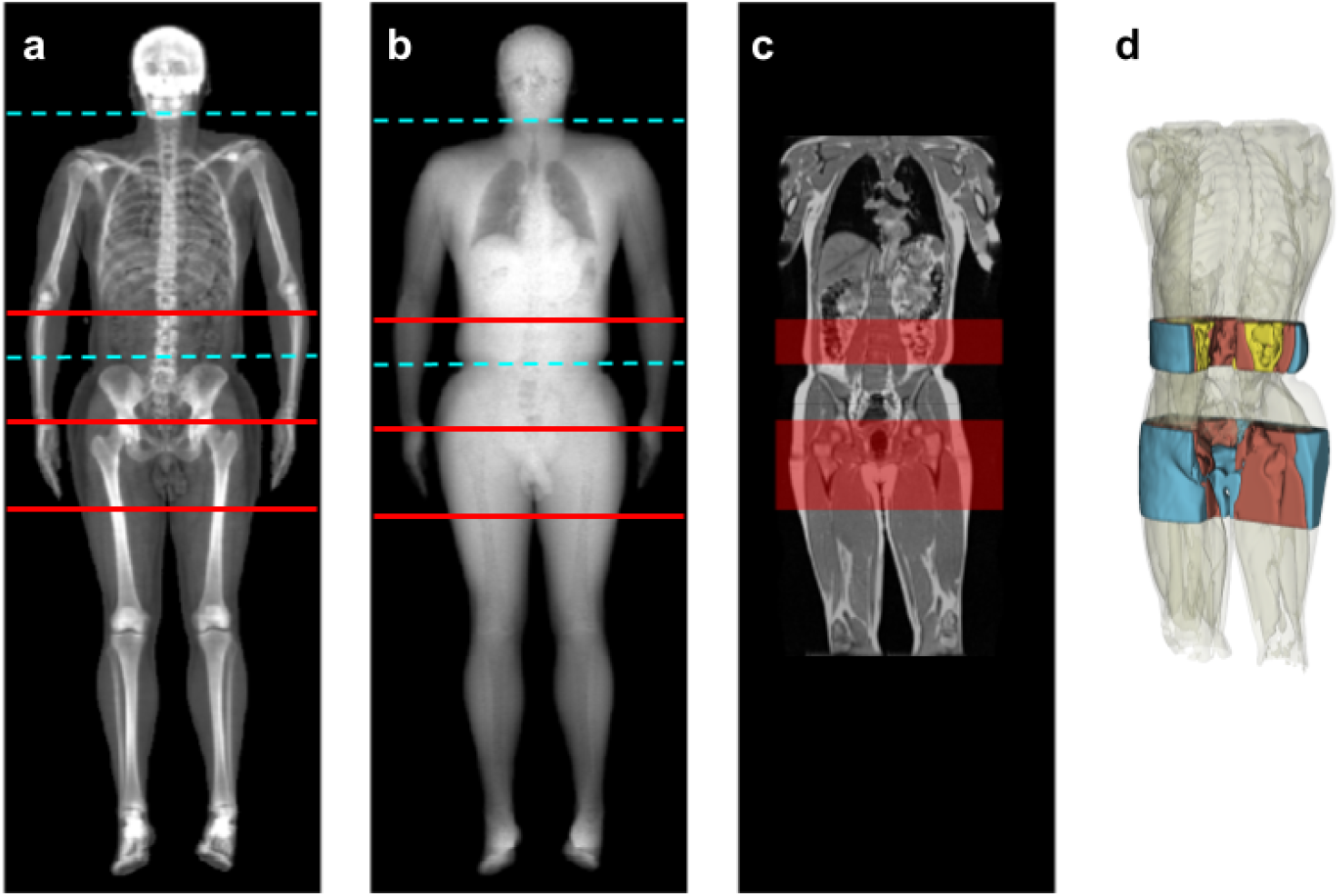
DXA and MRI scans with android and gynoid regions displayed. a) High-energy DXA with manually placed lines following the manufacturer’s definitions. Blue dashed lines are the neck and pelvic cuts. The android region spans from the first red line to the pelvic cut, and the gynoid region spans the area between the bottom two red lines. b) Low-energy DXA with the same cuts and areas. c) MRI of the same participant, with android and gynoid regions overlaid. d) 3D view of the MRI-based segmentations, with structures inside non-DXA regions greyed out. Blue = subcutaneous adipose tissue (SAT). Yellow = visceral adipose tissue (VAT). Red = skeletal muscle tissue.

Given the ability of MRI to differentiate between various tissues within any anatomical area of interest, we categorised “lean tissue” in the DXA-android region to include all non-fat tissues, such as muscle, kidney, and liver, as assessed by MRI. In the gynoid region, lean tissue was defined as all non-adipose and non-bone marrow tissues. To gain further insight, we conducted a detailed comparison between the “lean tissue” identified by DXA and the specific muscle volume measured by MRI. This comparison aimed to highlight any discrepancies between the broader classification of lean tissue in DXA and the more precise muscle measurements provided by MRI, helping us better understand the gradations between these two modalities in body composition assessment.

For the DXA measures that are given in mass, we converted MRI volumes to mass, using density values. Given the lack of consensus in the literature regarding the density of muscle vs lean body mass, we used 1.06g/ml for both^25–27^, 1.04g/ml for kidney^28,29^, and 1.05g/ml for liver^30,31^. Similarly, we took into account the fact that adipose tissue volume measured by MRI and fat mass measured by DXA are not equivalent. Indeed, it is accepted that around 80% of adipose tissue is fat^32^, and we adjusted adipose tissue volume measurements accordingly. Adipose tissue volume measured by MRI was adjusted to DXA fat mass using a fat density of 0.92g/ml^33,34^.

### Statistical Analysis

Descriptive characteristics are presented as means and standard deviations (SD) using the R software environment^35^. The Wilcoxon rank-sum test was used to compare means between groups, and the Wilcoxon signed-rank test was used to compare groups with paired observations. A linear regression model was fit to extract the regression equation between DXA and MRI measures. After counting the hypothesis tests, the Bonferroni-corrected threshold for statistical significance was 0.05/53 = 0.00094.

## Results

Baseline characteristics and summary statistics for the whole cohort are provided in supplementary Table S1. Among the 31,033 participants who attended the baseline imaging visit, 96.2% were White, and 47.5% were male. The participants had an average age of 64.1 ± 7.8 years and BMI of 26.5 ± 4.4 kg/m^2^.

Table 1 provides measurements obtained from DXA and MRI in the android and gynoid regions at the baseline imaging visit. These measurements include muscle mass, lean mass, fat tissue mass, and body fat percentage, separated by sex. The VAT volume was further assessed in the android region. Supplementary figures S1-S7, show the relationship between MRI and DXA measures for all participants at the baseline imaging visit.

**Table 1.** Body composition measured via DXA and MRI at the baseline imaging visit separated by sex within the android and gynoid regions. N_Android_ = 30,892. N_Gnyoid_ = 31,033. After Bonferroni correction (p = 0.00094), all linear regression coefficients were statistically significant.

| Android Region | Men (N = 14,996) |  |  |  | Women (N = 15,896) |  |  |  |
| --- | --- | --- | --- | --- | --- | --- | --- | --- |
|  | MRI | DXA | R<sup>2</sup> | Regression equation | MRI | DXA | R<sup>2</sup> | Regression equation |
| Muscle mass (kg) | 1.60 ± 0.28 | - | - | - | 0.98 ± 0.17 | - | - | - |
| Lean mass (kg) | 1.74 ± 0.31 | 4.10 ± 0.54 | 0.493 | y = 1.23x + 1.95 | 1.10 ± 0.20 | 2.92 ± 0.40 | 0.456 | y = 1.36x + 1.43 |
| Fat mass (kg) | 2.78 ± 1.11 | 2.62 ± 1.20 | 0.951 | y = 1.06x - 0.32 | 2.32 ± 1.06 | 2.15 ± 1.12 | 0.959 | y = 1.03x - 0.25 |
| Fat percentage (%) | 52.64 ± 11.17 | 37.33 ± 10.25 | not linear | not linear | 55.89 ± 12.56 | 40.03 ± 11.56 | not linear | not linear |
| Visceral adipose tissue volume (l) | 2.03 ± 0.98 | 1.77 ± 1.04 | 0.9 | y = 1.01x - 0.27 | 0.92 ± 0.57 | 0.79 ± 0.62 | 0.891 | y = 1.03x - 0.15 |
| Gynoid Region | Men (N = 15,037) |  |  |  | Women (N = 15,996) |  |  |  |
|  | MRI | DXA | R<sup>2</sup> | Regression equation | MRI | DXA | R<sup>2</sup> | Regression equation |
| Muscle mass (kg) | 6.02 ± 1.00 | - | - | - | 4.02 ± 0.66 | - | - | - |
| Lean mass (kg) | 7.78 ± 1.17 | 8.58 ± 1.11 | 0.749 | y = 0.82x + 2.22 | 5.33 ± 0.84 | 6.11 ± 0.79 | 0.726 | y = 0.8x + 1.84 |
| Fat mass (kg) | 3.68 ± 1.14 | 3.52 ± 1.23 | 0.949 | y = 1.05x - 0.33 | 4.62 ± 1.34 | 4.59 ± 1.52 | 0.925 | y = 1.09x - 0.46 |
| Fat percentage (%) | 38.44 ± 7.06 | 28.50 ± 6.06 | not linear | not linear | 53.78 ± 6.67 | 41.99 ± 6.63 | not linear | not linear |

The results show that DXA overestimates lean mass in the android region compared to MRI by approximately 1.95kg in men and 1.43kg in women (Table 1). Similarly, in the gynoid region, DXA overestimates MRI lean mass by 2.22kg in men and 1.84kg in women. The percent variance in DXA fat mass measures explained by the MRI fat mass measures (or R^2^ value) was 95.1% in men and 95.9% in women. Similar results were found for gynoid fat mass measurements. VAT also showed a good fit with R^2^ of 0.9 in both men and women. Similar results were observed in the follow-up imaging visit (Supplementary Table S2).

Table 2 provides a comparison of the longitudinal observations for MRI and DXA measurements, captured approximately 2.3 ± 0.1 years apart between the baseline and follow-up imaging visits. We observed a reduction in muscle and lean mass as measured by MRI in both android and gynoid regions for both sexes, with changes ranging from -4.5% to -3.9% in the angroid region and -5.2% to -4.1% in the gynoid region. In contrast, DXA lean mass in women showed a significant increase of 2.6%. This could be due to differences in anatomy captured within the android region caused by variations in breath-hold during imaging. Figure S8 shows an example of lean mass as measured by MRI in the android region including abdominal organs and muscle, at baseline and follow-up imaging for one participant, illustrating a 12.4% decrease in lean tissue when all organs are included, but only a 1% decrease when the liver and kidneys are omitted. No further significant changes were detected in lean mass measurements from DXA. We found no significant changes in any other tissue measurements from MRI or DXA.

**Table 2.** Longitudinal measures of body composition from DXA and MRI separated by sex within the android and gynoid regions. Significance refers to the p-value for a Wilcoxon rank-sums test, where the null hypothesis is that the medians between the two groups (baseline and follow-up imaging visits for MRI and DXA measures) are equal. An asterisk (*) indicates statistical significance for p-value < 0.05, ** indicates statistical significance after Bonferroni correction (p = 0.00094).

| Android Region | MRI |  |  |  | DXA |  |  |  |
| --- | --- | --- | --- | --- | --- | --- | --- | --- |
| Imaging Visit | Baseline | Follow-up | Difference | % Difference | Baseline | Follow-up | Difference | % Difference |
|  | **Men (N = 749)** |  |  |  |  |  |  |  |
| Muscle mass (kg) | 1.69 ± 0.28 | 1.61 ± 0.28** | -0.08 ± 0.15 | -4.40 ± 8.54 | - | - | - | - |
| Lean mass (kg) | 1.85 ± 0.31 | 1.76 ± 0.30** | -0.09 ± 0.19 | -4.35 ± 10.15 | 4.05 ± 0.52 | 4.11 ± 0.53* | 0.06 ± 0.21 | 1.64 ± 5.12 |
| Fat mass (kg) | 2.76 ± 1.16 | 2.70 ± 1.14 | -0.06 ± 0.49 | -0.87 ± 18.43 | 2.51 ± 1.23 | 2.57 ± 1.26 | 0.06 ± 0.50 | 4.51 ± 21.07 |
| Fat percentage (%) | 51.19 ± 11.36 | 51.64 ± 11.49 | 0.45 ± 4.92 | 1.49 ± 11.27 | 36.40 ± 10.40 | 36.60 ± 10.54 | 0.20 ± 4.56 | 1.47 ± 14.30 |
| Visceral adipose tissue volume (l) | 1.98 ± 1.00 | 1.96 ± 0.99 | -0.02 ± 0.42 | 1.55 ± 23.09 | 1.63 ± 1.06 | 1.72 ± 1.09 | 0.09 ± 0.41 | 11.11 ± 30.28 |
|  | **Women (N = 665)** |  |  |  |  |  |  |  |
| Muscle mass (kg) | 1.02 ± 0.17 | 0.97 ± 0.16** | -0.05 ± 0.09 | -4.51 ± 8.46 | - | - | - | - |
| Lean mass (kg) | 1.14 ± 0.19 | 1.09 ± 0.23** | -0.05 ± 0.18 | -3.86 ± 14.15 | 2.83 ± 0.36 | 2.90 ± 0.35** | 0.07 ± 0.16 | 2.59 ± 5.63 |
| Fat mass (kg) | 2.23 ± 0.96 | 2.20 ± 0.97 | -0.03 ± 0.38 | -0.05 ± 18.27 | 1.97 ± 0.98 | 2.05 ± 1.03 | 0.09 ± 0.38 | 5.90 ± 19.98 |
| Fat percentage (%) | 54.35 ± 12.53 | 54.98 ± 12.70 | 0.63 ± 4.51 | 1.60 ± 10.13 | 38.79 ± 11.20 | 39.15 ± 11.47 | 0.36 ± 4.33 | 1.63 ± 13.01 |
| Visceral adipose tissue volume (l) | 1.00 ± 0.55 | 1.01 ± 0.57 | 0.01 ± 0.21 | 3.16 ± 25.51 | 0.69 ± 0.55 | 0.75 ± 0.59 | 0.06 ± 0.21 | 47.38 ± 288.6 |
| Gynoid Region | MRI |  |  |  | DXA |  |  |  |
| Imaging Visit | Baseline | Follow-up | Difference | % Difference | Baseline | Follow-up | Difference | % Difference |
|  | **Men (N = 750)** |  |  |  |  |  |  |  |
| Muscle mass (kg) | 6.38 ± 0.98 | 6.05 ± 0.97** | -0.33 ± 0.59 | -4.81 ± 9.14 | - | - | - | - |
| Lean mass (kg) | 8.15 ± 1.15 | 7.79 ± 1.13** | -0.36 ± 0.68 | -4.09 ± 8.36 | 8.60 ± 1.07 | 8.56 ± 1.08 | -0.04 ± 0.30 | -0.42 ± 3.45 |
| Fat mass (kg) | 3.70 ± 1.19 | 3.63 ± 1.16 | -0.07 ± 0.43 | -1.28 ± 11.37 | 3.46 ± 1.24 | 3.48 ± 1.24 | 0.02 ± 0.44 | 1.43 ± 12.35 |
| Fat percentage (%) | 37.40 ± 7.08 | 37.93 ± 7.07 | 0.53 ± 2.56 | 1.72 ± 7.40 | 28.07 ± 6.00 | 28.30 ± 6.03 | 0.23 ± 2.40 | 1.21 ± 8.82 |
|  | **Women (N = 689)** |  |  |  |  |  |  |  |
| Muscle mass (kg) | 4.20 ± 0.65 | 3.97 ± 0.61** | -0.24 ± 0.38 | -5.19 ± 8.75 | 6.08 ± 0.72 | 6.02 ± 0.74 | -0.05 ± 0.33 | -0.78 ± 5.17 |
| Lean mass (kg) | 5.51 ± 0.79 | 5.26 ± 0.84** | -0.25 ± 0.57 | -4.24 ± 9.75 | 6.08 ± 0.72 | 6.02 ± 0.74 | -0.05 ± 0.33 | -0.78 ± 5.17 |
| Fat mass (kg) | 4.64 ± 1.24 | 4.49 ± 1.26* | -0.15 ± 0.53 | -2.90 ± 11.09 | 4.42 ± 1.37 | 4.48 ± 1.47 | 0.06 ± 0.58 | 1.75 ± 12.17 |
| Fat percentage (%) | 53.15 ± 6.54 | 53.41 ± 6.70 | 0.25 ± 2.50 | 0.56 ± 4.86 | 41.29 ± 6.43 | 41.68 ± 6.78 | 0.39 ± 3.22 | 1.17 ± 7.62 |

## Discussion

In this study, we compared MRI measurements of muscle, lean and adipose tissue mass, and body fat percentage in the android and gynoid regions—areas typically assessed in DXA scans—to DXA measurement estimates in these regions. We assessed the linear relationship between the MRI and DXA-derived body composition measurements in the android and gynoid regions, and we assessed the longitudinal changes in body composition measures by DXA and MRI during follow-up imaging visits. Our primary aim was to evaluate the agreement between MRI and DXA measurements in anatomically equivalent regions and to determine their precision in detecting longitudinal changes.

Android and gynoid regions are constructs specifically defined in DXA scans. Our study replicated these constructs in our MRI measurements to allow a direct comparison between these two imaging methods. DXA showed a good agreement against MRI for adipose tissue measures, with a strong correlation between MRI and DXA fat mass measures, aligning with previous literature^36^. However, in our cross-sectional analysis we found that DXA overestimated lean mass when compared to MRI in both android and gynoid regions. Although previous studies comparing MRI-derived muscle against DXA-derived lean mass, found a strong correlation between the two^17,37,38^, others comparing CT and DXA, have reported that DXA overestimates muscle, with larger differences in subjects with higher body weights, suggesting DXA may be less accurate in that population^7^.

Longitudinal assessment of body composition is vitally important, particularly in ageing, where a decline in skeletal muscle mass, and loss of muscle strength^39^ contribute to sarcopenia and progression toward frailty^40^. We found that over the relatively short period of 2.5 years, both muscle and lean mass, as measured by MRI, decreased by 4% to 5% in both android and gynoid regions. This decrease in both muscle and lean mass after two years, likely reflects both a loss of muscle and an increase in intramuscular fat content in that period, both characteristic of the ageing process. The apparent lack of change observed by DXA may reflect its inability to differentiate between these compartments. Previous longitudinal studies in older adults have reported that MRI and DXA detected a similar percentage loss of thigh muscle mass over a five-year period^13^, with similar agreement reported for changes in leg skeletal muscle mass following two years exercise training^38^. However, several interventional studies have suggested that compared with MRI and other gold standard methods, DXA is unable to accurately detect longitudinal changes in lean muscle^17,41^. This inability is clearly reflected in the current longitudinal study.

Unexpectedly, our study found a significant increase in DXA-measured lean mass in the android region among women at follow-up. The reason for this increase, or why it was observed only in women, remains unclear. Prior research has shown that DXA measurements of lean soft tissue mass in the trunk can differ from MRI measurements of muscle in the same region^38^. This difference may result from DXA’s limited capacity to distinguish muscle from other lean tissues with differing proteins, water, non-fat lipids, carbohydrates, and soft tissue minerals composition^6^. Additionally, changes in fluid retention could further complicate these measurements, potentially contributing to the inconsistencies observed.

Our study was not without limitations, particularly regarding the variability in the neck placement for the MRI scan, which is essential for our android ROI height definition. However, the impact of this variation is mitigated somewhat since the android ROI height is set only to be one-fifth of the distance between the neck and the pelvic cut. For longitudinal datasets, we fixed the height of the ROIs to match baseline images in order to remove any variability from neck positioning. Furthermore, although DXA appendicular lean mass is a more commonly used measure of muscle^42^, we are unable to directly compare these measures in our study since the UK Biobank neck-to-knee MRI acquisition does not include the arms and lower legs. Finally, the interval between baseline and follow-up measurements in the longitudinal aspect of this study was only two and a half years, and the changes detected by MRI although consistent were modest^24^. Longer intervals between scans or interventions designed to produce significant weight change may result in more significant changes in lean mass which may be more reliably detected by DXA.

## Conclusion

Our findings in this large-scale study underscore the limitations and strengths of DXA for body composition analysis. While DXA provides reliable cross-sectional estimates for both fat and VAT, it overestimates lean mass and has considerable limitations for the assessment of longitudinal changes; an essential consideration for studies into ageing. The more detailed body composition assessments and precise measurements from MRI are therefore more appropriate for understanding health in longitudinal population studies.

## Acknowledgements

This study was carried out using UK Biobank Application number 44584, and we thank the participants in the UK Biobank imaging study. This study was funded by Calico Life Sciences LLC.

## Author Contributions

J.D.B., E.L.T., M.T and N.B. conceived the study. All authors designed the study. M.T., B.W. and N.B implemented the methods. N.B. performed the data analysis and M.T. performed the statistical analysis. All authors drafted the manuscript. All authors read and approved the manuscript.

## Declarations

Participant data from the UK Biobank cohort was obtained through UK Biobank Access Application number 44584. The UK Biobank has approval from the North West Multi-Centre Research Ethics Committee (REC reference: 11/NW/0382), with informed consent obtained from all participants. Researchers may apply to use the UKBB data resource by submitting a health-related research proposal in the public interest. Additional information may be found on the UK Biobank researchers and resource catalogue pages (http://www.ukbiobank.ac.uk). All methods were performed in accordance with the relevant guidelines and regulations as presented by the appropriate authorities, including the Declaration of Helsinki.

## Data Availability

Our research was conducted using UK Biobank data. Under the standard UK Biobank data sharing agreement, we (and other researchers) cannot directly share raw data obtained or derived from the UK Biobank. However, under this agreement, all of the data generated, and methodologies used in this paper are returned by us to the UK Biobank, where they will be fully available. Access is obtained directly from the UK Biobank to all bona fide researchers upon submitting a health-related research proposal to the UK Biobank https://www.ukbiobank.ac.uk.

## Footnotes

1 https://biobank.ndph.ox.ac.uk/showcase/label.cgi?id=124

2 X-ray bone densitometer with enCORE v17 software user manual https://www.gehealthcare.com/support/manuals

