## Supplementary material for "Comparing DXA and MRI body composition measurements in cross-sectional and longitudinal cohorts"

### Comparing DXA to MRI for body composition measurements at population scale - Supplementary Material

Supplementary Tables

| | Baseline imaging visit | | | Follow-up imaging visit | | |
|---|---|---|---|---|---|---|
| | **Full Cohort (N = 31,033)** | **Men (N = 14,743)** | **Women (N = 15,711)** | **Full Cohort (N = 1,928)** | **Men (N = 993)** | **Women (N = 935)** |
| **White (n)** | 29,858 | 14,419 | 15,306 | 1,875 | 961 | 881 |
| **Age (yrs.)** | 64.17 ± 7.75 | 64.88 ± 7.84 | 63.52 ± 7.61 | 65.21 ± 7.22 | 65.68 ± 7.20 | 64.80 ± 7.17 |
| **Weight (kg)** | 76.00 ± 15.09 | 83.48 ± 13.13 | 68.71 ± 12.73 | 75.57 ± 14.69 | 82.54 ± 12.91 | 67.44 ± 11.90 |
| **Height (m)** | 169.23 ± 9.29 | 176.12 ± 6.66 | 162.75 ± 6.26 | 169.82 ± 9.46 | 176.50 ± 6.80 | 162.68 ± 6.13 |
| **BMI (kg/m²)** | 26.45 ± 4.35 | 26.89 ± 3.82 | 25.94 ± 4.61 | 26.12 ± 4.21 | 26.48 ± 3.76 | 25.48 ± 4.31 |
| **Waist circumference (cm)** | 87.65 ± 12.62 | 93.61 ± 10.46 | 81.85 ± 11.51 | 87.93 ± 12.08 | 93.15 ± 10.29 | 81.81 ± 10.78 |
| **Hip circumference (cm)** | 100.37 ± 8.67 | 100.25 ± 7.17 | 100.33 ± 9.63 | 99.57 ± 8.17 | 99.39 ± 7.00 | 99.23 ± 8.69 |
| **WHR** | 0.87 ± 0.09 | 0.93 ± 0.06 | 0.81 ± 0.07 | 0.88 ± 0.09 | 0.94 ± 0.06 | 0.82 ± 0.07 |
| **Systolic blood pressure (mmHg)** | 138.35 ± 18.57 | 141.51 ± 17.35 | 135.34 ± 19.17 | 139.93 ± 17.99 | 142.58 ± 16.87 | 136.99 ± 18.56 |
| **Diastolic blood pressure (mmHg)** | 78.57 ± 10.03 | 80.41 ± 9.80 | 76.80 ± 9.93 | 78.85 ± 9.78 | 80.71 ± 9.74 | 76.85 ± 9.46 |
| **Dominant hand grip strength (kg)** | 31.13 ± 10.63 | 38.73 ± 8.99 | 23.94 ± 6.07 | 28.40 ± 10.58 | 35.21 ± 9.24 | 21.08 ± 6.12 |

Table S1: Demographics of the full cohort, separated by sex. Values are reported as mean ± standard deviation for continuous variables and counts (N) for categorical variables.

| | Men (N = 992) | | | | Women (N = 903) | | | |
|---|---|---|---|---|---|---|---|---|
| **Android Region** | **MRI** | **DXA** | **R²** | **Regression equation** | **MRI** | **DXA** | **R²** | **Regression equation** |
| **Muscle mass (kg)** | 1.60 ± 0.27 | - | - | - | 0.97 ± 0.16 | - | - | - |
| **Lean mass (kg)** | 1.75 ± 0.30 | 4.11 ± 0.53 | 0.542 | y = 1.3x + 1.85 | 1.09 ± 0.22 | 2.92 ± 0.38 | 0.375 | y = 1.05x + 1.78 |
| **Fat mass (kg)** | 2.70 ± 1.10 | 2.57 ± 1.21 | 0.956 | y = 1.07x - 0.33 | 2.27 ± 1.01 | 2.12 ± 1.09 | 0.966 | y = 1.06x - 0.28 |
| **Fat percentage (%)** | 51.93 ± 11.32 | 36.70 ± 10.35 | not linear | not linear | 55.61 ± 12.53 | 39.67 ± 11.50 | not linear | not linear |
| **Visceral adipose tissue volume (l)** | 1.97 ± 0.96 | 1.73 ± 1.05 | 0.908 | y = 1.04x - 0.32 | 1.04 ± 0.59 | 0.78 ± 0.62 | 0.89 | y = 0.98x - 0.24 |
| | **Men (N = 993)** | | | | **Women (N = 935)** | | | |
| **Gynoid Region** | **MRI** | **DXA** | **R²** | **Regression equation** | **MRI** | **DXA** | **R²** | **Regression equation** |
| **Muscle mass (kg)** | 6.02 ± 0.96 | - | - | - | 3.96 ± 0.61 | - | - | - |
| **Lean mass (kg)** | 7.76 ± 1.12 | 8.55 ± 1.07 | 0.781 | y = 0.84x + 2.01 | 5.27 ± 0.84 | 6.06 ± 0.76 | 0.61 | y = 0.7x + 2.37 |
| **Fat mass (kg)** | 3.61 ± 1.12 | 3.46 ± 1.19 | 0.96 | y = 1.04x - 0.3 | 4.52 ± 1.30 | 4.54 ± 1.54 | 0.921 | y = 1.13x - 0.59 |
| **Fat percentage (%)** | 37.97 ± 7.00 | 28.26 ± 5.98 | not linear | not linear | 53.56 ± 6.61 | 41.87 ± 6.76 | not linear | not linear |

Table S2: Body composition measured via DXA and MRI at the follow-up imaging visit separated by sex within android and gynoid regions. $N_{Android}$ = 1,895. $N_{Gnyoid}$ = 1,928. Android muscle mass was not available from DXA measures, so the regression was done between DXA lean mass and MRI muscle mass. After Bonferroni correction (p = 0.00094), all the linear regression coefficients were statistically significant.

#### Supplementary Figures

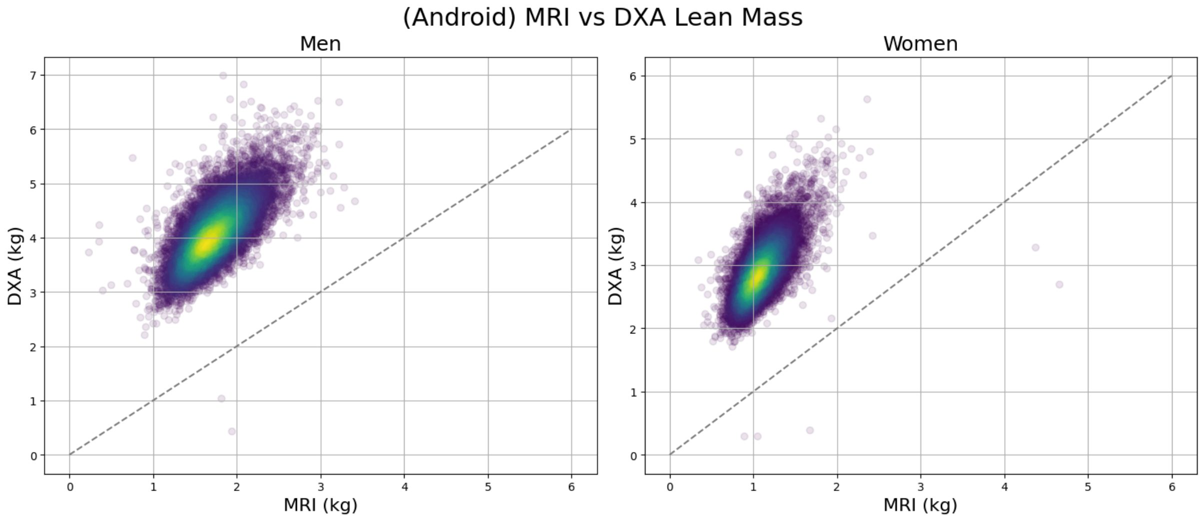


Figure S1: Scatterplots of DXA (y-axis) by MRI (x-axis) android lean mass (kg), separated by sex.

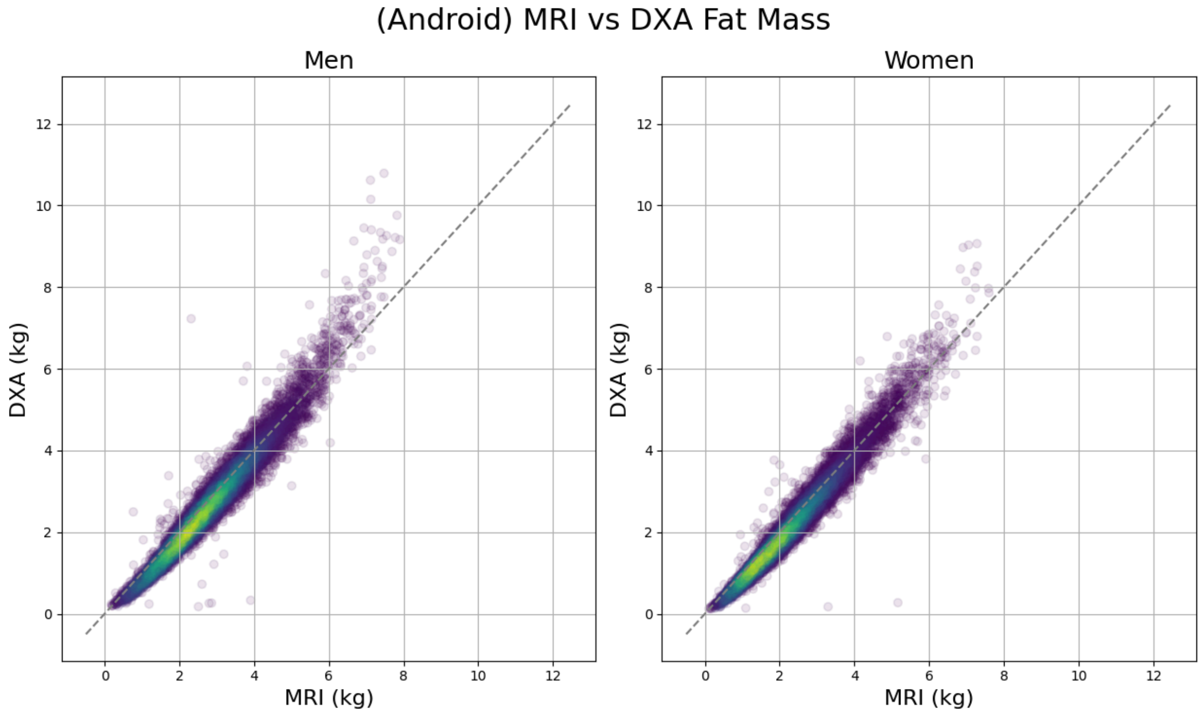


Figure S2: Scatterplots of DXA (y-axis) by MRI (x-axis) android fat mass (kg), separated by sex.

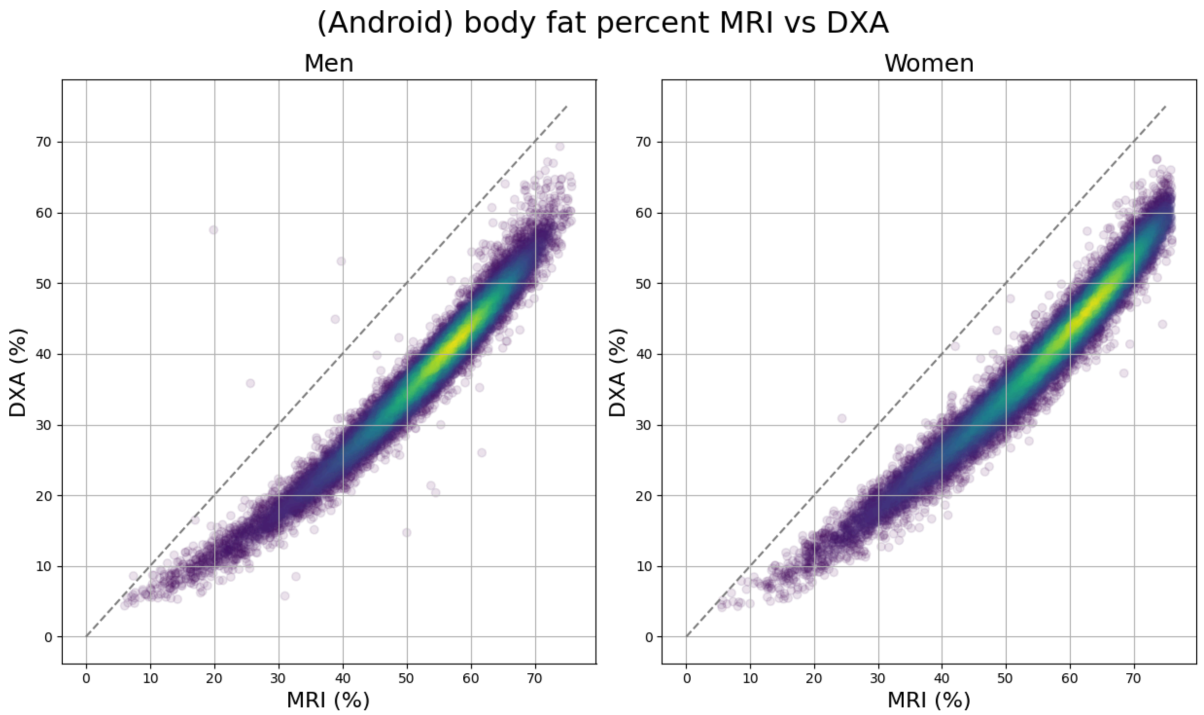


Figure S3: Scatterplots of DXA (y-axis) by MRI (x-axis) android fat percent (%), separated by sex.

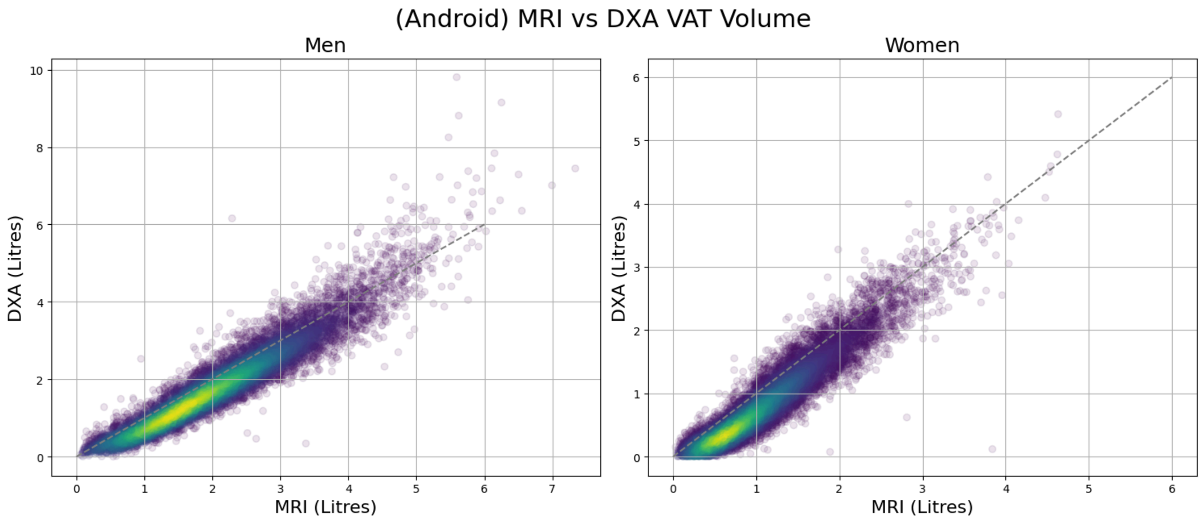


Figure S4: Scatterplots of DXA (y-axis) by MRI (x-axis) android visceral adipose tissue (VAT) volume (litres), separated by sex.

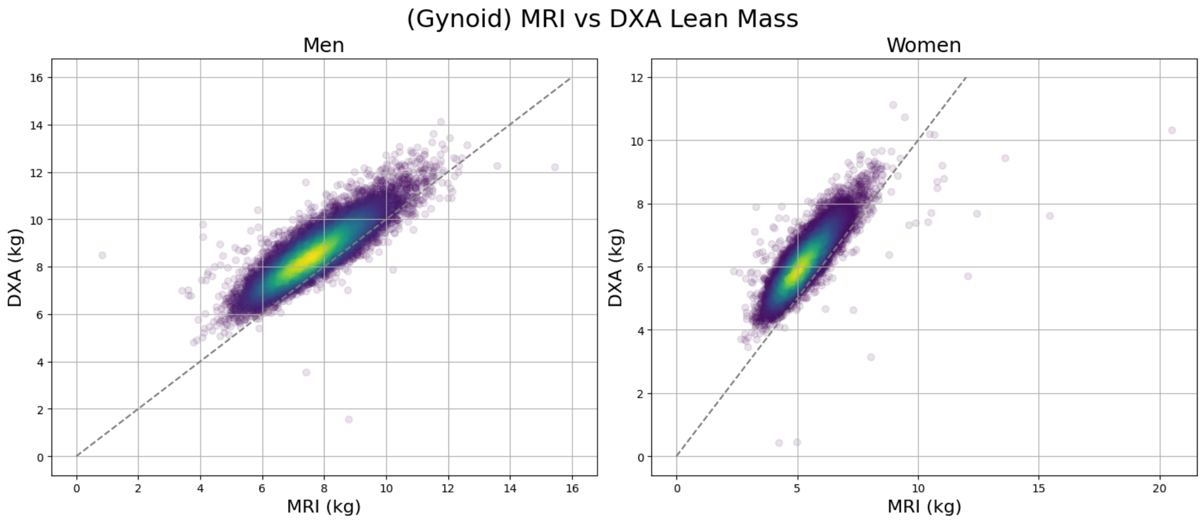


Figure S5: Scatterplots of DXA (y-axis) by MRI (x-axis) gynoid lean mass (kg), separated by sex.

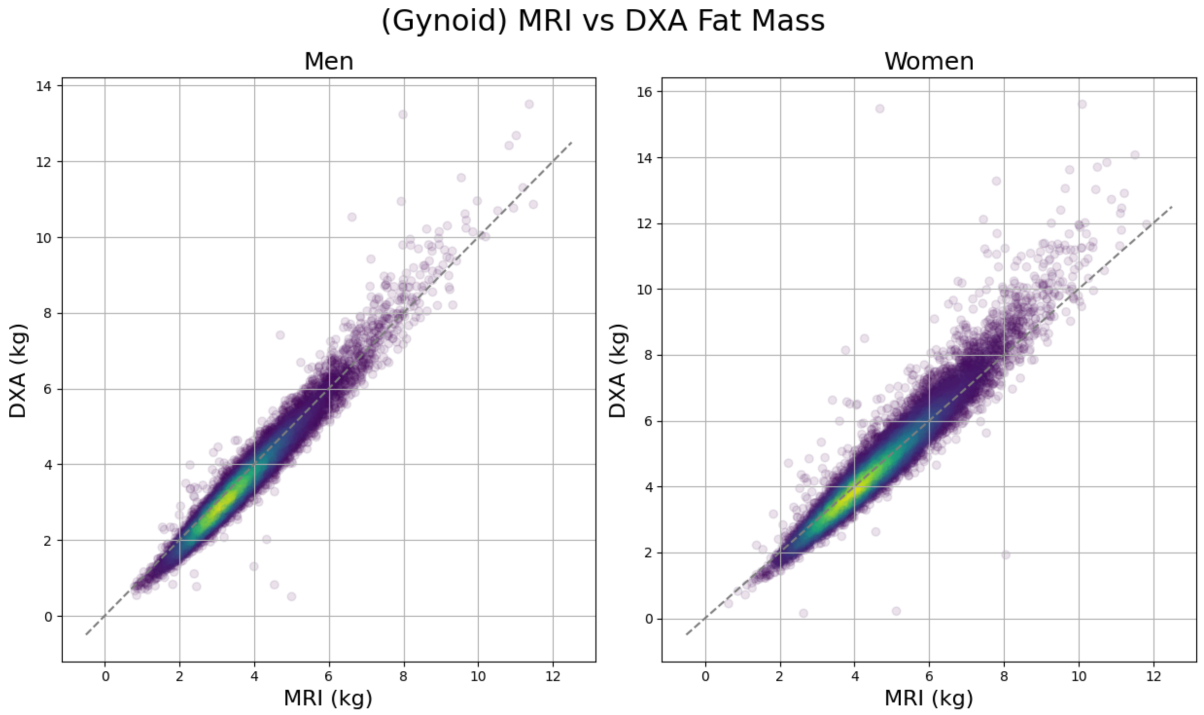


Figure S6: Scatterplots of DXA (y-axis) by MRI (x-axis) gynoid fat mass (kg), separated by sex.

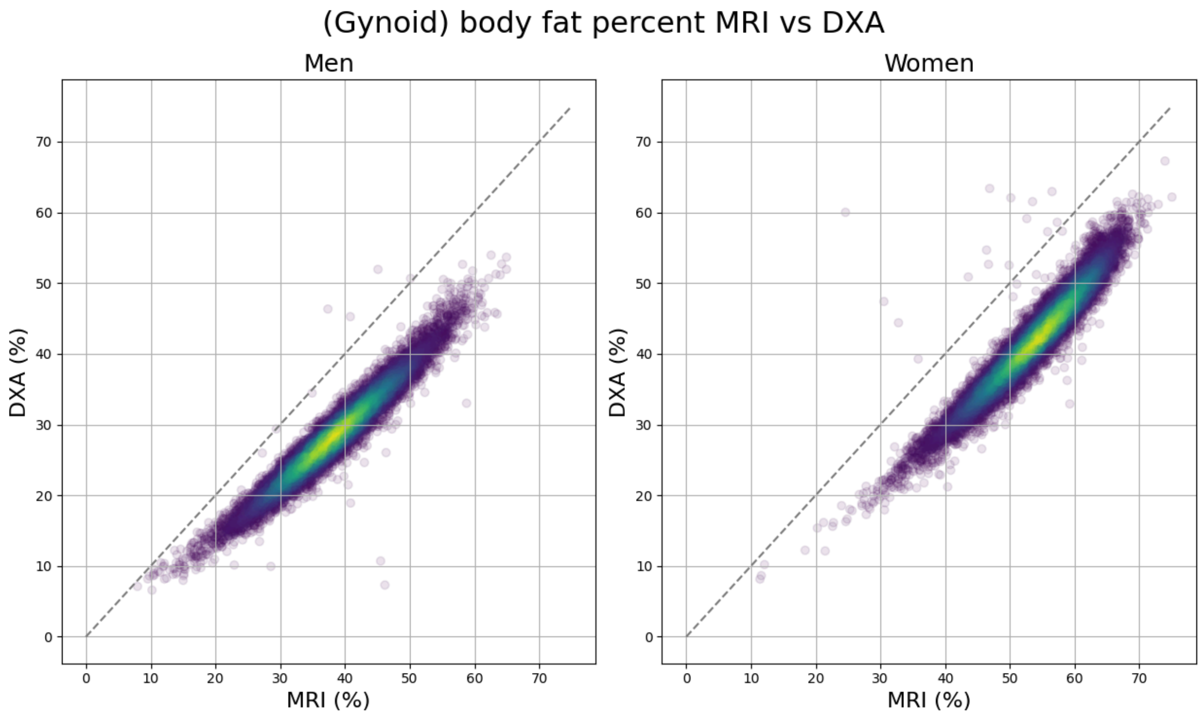


Figure S7: Scatterplots of DXA (y-axis) by MRI (x-axis) gynoid fat percentage (%), separated by sex.

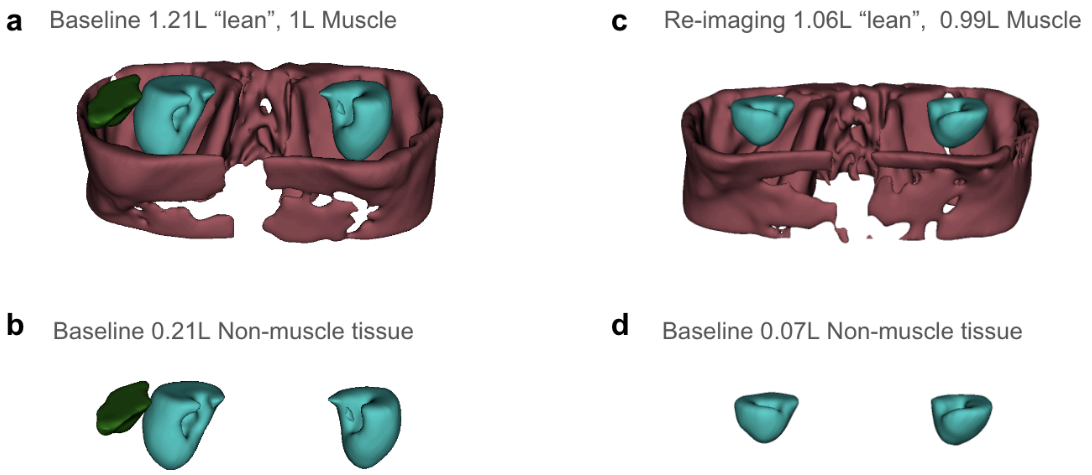


Figure S8: Example of MRI android lean tissue including abdominal organs and muscle at baseline (a,b) and follow-up (c,d) imaging visit for one participant. The figure demonstrates a 12.4% decrease in lean tissue when including the liver and kidneys, but only a 1% decrease when these organs are omitted, highlighting the impact of different breath holds on anatomy captured within the android region.
